# Risk estimation and prediction by modeling the transmission of the novel coronavirus (COVID-19) in mainland China excluding Hubei province

**DOI:** 10.1101/2020.03.01.20029629

**Authors:** Hui Wan, Jing-an Cui, Guo-jing Yang

## Abstract

**Background:** In December 2019, an outbreak of coronavirus disease (COVID-19) was identified in Wuhan, China and, later on, detected in other parts of China. Our aim is to evaluate the effectiveness of the evolution of interventions and self-protection measures, estimate the risk of partial lifting control measures and predict the epidemic trend of the virus in mainland China excluding Hubei province based on the published data and a novel mathematical model.

**Methods:** A novel COVID-19 transmission dynamic model incorporating the intervention measures implemented in China is proposed. COVID-19 daily data of mainland China excluding Hubei province, including the cumulative confirmed cases, the cumulative deaths, newly confirmed cases and the cumulative recovered cases for the period January 20th-March 3rd, 2020, were archived from the National Health Commission of China (NHCC). We parameterize the model by using the Markov Chain Monte Carlo (MCMC) method and estimate the control reproduction number *R*_*c*_, as well as the effective daily reproduction ratio *R*_*e*_(*t*), of the disease transmission in mainland China excluding Hubei province.

**Results:** The estimation outcomes indicate that *R*_*c*_ is 3.36 (95% CI 3.20-3.64) and *R*_*e*_(*t*) has dropped below 1 since January 31st, 2020, which implies that the containment strategies implemented by the Chinese government in mainland China excluding Hubei province are indeed effective and magnificently suppressed COVID-19 transmission. Moreover, our results show that relieving personal protection too early may lead to the spread of disease for a longer time and more people would be infected, and may even cause epidemic or outbreak again. By calculating the effective reproduction ratio, we prove that the contact rate should be kept at least less than 30% of the normal level by April, 2020.

**Conclusions:** To ensure the epidemic ending rapidly, it is necessary to maintain the current integrated restrict interventions and self-protection measures, including travel restriction, quarantine of entry, contact tracing followed by quarantine and isolation and reduction of contact, like wearing masks, etc. People should be fully aware of the real-time epidemic situation and keep sufficient personal protection until April. If all the above conditions are met, the outbreak is expected to be ended by April in mainland China apart from Hubei province.

## Background

Coronaviruses are a group of enveloped viruses with a positive-sense, single-stranded RNA and viral particles resembling a crown, from which the name derives. They belong to the order of *Nidovirales*, family of *Coronaviridae*, and subfamily of *Orthocoronavirinae* ([1]).

In December 2019, an outbreak of coronavirus disease (COVID-19) was identified in Wuhan, China and, later on, detected in other parts of China. By February 27th, the new virus had infected more than 78,900 people and killed at least 2,791 in China ([2]). Besides China, more than 4,440 people had been infected and 67 died in at least 48 countries and regions ([3]). Currently, there exist no vaccines or anti-viral treatments officially approved for the prevention or management of the diseases. The outbreaks are still on-going.

The basic reproduction number *R*_0_ is the average number of secondary infections due to an infective during the infectious period when everyone else in the population is susceptible [22]. While the basic reproduction number with control measures is defined as the control reproduction number *R*_*c*_. At the early stage of the outbreak, estimation of *R*_0_/*R*_*c*_ is crucial for determining the potential and severity of an outbreak, and providing precise information for designing and implementing disease outbreak responses, namely the identification of the most appropriate, evidence-based interventions, mitigation measures and the determination of the intensity of such programs in order to achieve the maximal protection of the population with the minimal interruption of social-economic activities [4, 5].

Recently, some papers have been released as pre-prints or undergone peer-review and published to estimate *R*_0_ and the risk of outbreak. Li et al. ([6]) analyzed data on the first 425 confirmed cases in Wuhan and determined the epidemiologic characteristics of COVID-19. Based on their estimates, the mean incubation period was 5.2 days, and *R*_0_ was 2.2, which is in line with the result estimated by Riou et al. ([7]). Zhao et al. ([8]) assessed the unreported number of COVID-19 cases in China in the first half of January with the estimation of *R*_0_ 2.56. Considering the impact of the variations in disease reporting rate, Zhao et al. ([9]) modelled the epidemic curve of COVID-19 cases, in mainland China from January 10 to January 24, 2020, through the exponential growth and concluded that the mean *R*_0_ ranged from 2.24 to 3.58 associated with 8-fold to 2-fold increase in the reporting rate. Li et al. ([10]) conducted a mathematical modeling study using five independent methods to assess *R*_0_ of COVID-19. Their results illustrated that *R*_0_ dropped from 4.38 to 3.41 after the closure of Wuhan city. Over the entire epidemic period COVID-19 had a *R*_0_ of 3.39. Moreover, Tang et al. formulated a deterministic compartmental model. Their estimations based on likelihood and model analysis showed that *R*_0_ with control measures might be as high as 6.47 ([5]). Most recently, Chen et al. ([11]) developed a Bats-Hosts-Reservoir-People transmission network model to simulate the potential transmission from the infectious sources to human. The estimated values of *R*_0_ were 2.30 from reservoir to person and 3.58 from person to person. We noticed that the estimations of *R*_0_ in varied studies are different. As mentioned in [5, 12], variability in the estimation of the basic reproduction number is a general recognized methodological issue, and standardized methods both for calculating and reporting *R*_0_ are still missing. Furthermore, the value of *R*_0_ may vary with key clinical parameters inferred from data which depend on the time period, quality, accuracy, and reliability. To better quantify the evolution of the interventions, Tang et al. fitted the previously proposed model in [5] to the data available until January 29th, 2020 and re-estimated the effective daily reproduction ([13]). There are also some literatures focusing on the prediction of COVID-19 development trend. Wang et al. formulated a complex network model and analyzed the possible time node and the risk impact of resumption on secondary outbreak in Wuhan and surrounding areas ([14]). Roosa et al. ([15]) utilized several dynamic models to forecast the cumulative number of confirmed cases in the coming 5, 10, and 15 days in Hubei province, and the overall trajectory of the epidemic in China excluding Hubei.

With the gradual alleviation of the epidemic situation in mainland China excluding Hubei province, considering the pressure of economic operation and the needs of people’s normal production and life, some places have adjusted the primary response of epidemic prevention and control to the secondary response [16]. In this situation, some critical questions need to be answered promptly. Does the reduction of the emergency response level mean that people can fully or partially relieve from self-production? When can people return back to normal life?

The aim of this work is to evaluate the effectiveness of varied interventions and self-protection measures, estimate the risk of partial lifting control measures and predict the epidemic trend of the virus in mainland China excluding Hubei province by establishing a COVID-19 transmission model incorporating the intervention measures implemented and fitting the data obtained from the National Health Commission of China (NHCC).

## Methods

### Establishment of COVID-19 transmission dynamic model

Based on the clinical progression of the disease, epidemiological status of the individuals and intervention measures (including travel restriction, body temperature measurement, close contact tracing, self-isolation and protection, etc.), we propose a novel deterministic COVID-19 transmission model. We parameterize the model using data of mainland China excluding Hubei province obtained from NHCC, and estimate the control reproduction number as well as the effective daily reproduction ratio of the disease transmission.

The population was grouped into various compartments, namely susceptible (S), exposed (E), infectious with symptoms (I), infectious but asymptomatic (A), isolated susceptible (Si), quarantined infected pending for confirmation (Q), hospitalized (H), and recovered (R). We assume that recovered individuals have immunity at least during this epidemic period. Let *N* (*t*) = *S*(*t*) + *E*(*t*) + *I*(*t*) + *A*(*t*) + *R*(*t*) be the total number of individuals in the free community. In order to fit the data, we explicitly generated additional two groups, i.e. the cumulative number of recovered *R*_*h*_(*t*) and dead cases *D*(*t*) from hospital. The total number of cumulative reported cases is set to be *T* (*t*). All the state variables are summarized in Table 1.

**Table 1.** State variables and initial values in Model (1).

| Definitions | State variables | Initial values | Geweke | Source |
| --- | --- | --- | --- | --- |
| Susceptible population | $S(t)$ | 1,336,210,000 | – | [27] |
| Exposed population | $E(t)$ | 501.23 | 0.92956 | MCMC |
| Symptomatic infected population | $I(t)$ | 0.22839 | 0.95511 | MCMC |
| Asymptomatic infected population | $A(t)$ | 991.29 | 0.81969 | MCMC |
| Isolated susceptible population | $S_i(t)$ | 0 | – | Data |
| Quarantined infected population | $Q(t)$ | 0 | – | Data |
| Infected population in hospitals | $H(t)$ | 21 | – | Data |
| Recovered population | $R(t)$ | 240.76 | 0.63851 | MCMC |
| Recovered population from hospitals | $R_h(t)$ | 0 | – | Data |
| Cumulative number of dead cases in hospitals | $D(t)$ | 0 | – | Data |
| Total number of reported cases | $T(t)$ | 21 | – | Data |

Due to the travel restriction, migrations from/to Hubei province and other regions are ignored. Birth and nature death are also neglected. With the increase of the cumulative number of confirmed cases, the probability of contact transmission among the informed susceptible populations would certainly reduce ([17, 18, 19], etc.). To better quantify the varied interventions and self-protection measures, we assume the contact rate to be time-dependent *c*(*t*) = *q*_1_(*t*)*c*_0_, where *c*_0_ is the initial contact rate and *q*_1_(*t*) is the intervention coefficient with respect to contact. Here we assume that *q*_1_(*t*) = *e*^*−δT* (*t*)^, which is dependent on the total number of cumulative confirmed cases *T* (*t*) and is monotone decreasing with *T* (*t*), so as to well reflect the impact of media coverage on people’s psychology and behaviors. *c*(*t*) = *c*_0_ for *T* = 0 and 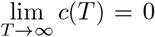. It should be mentioned that the contact function *c*(*t*) in [13] is also assumed to be time-dependent, but it is not dependent on state variables. Let the transmission probability be *β*. Thus, the incidence rate can be given by 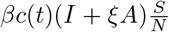, where *ξ* is the correction factor of transmission probability with asymptomatic infectious individuals.

By body temperature measurements everywhere and diagnoses in hospitals, symptomatic infectious individuals can be detected. The detection rate is assumed to be *q*_2_*I*(*t*). Infected individuals in *Q* class can be confirmed at the rate of *ηQ*(*t*) by nucleic acid testing. Additionally, close contact tracing followed by quarantine and isolation is a critical control measure. We assume that, once a case is confirmed, *q*_3_ individuals would be traced. Therefore, *q*_3_(*q*_2_*I*(*t*) + *ηQ*(*t*)) individuals would be traced in a unit time, which is dependent on the number of new confirmed cases *q*_2_*I*(*t*) + *ηQ*(*t*)). We also assume that, among these traced individuals, 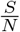 fraction parts are susceptible, 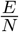 fraction parts are exposed, 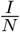 fraction parts are infectious with symptoms and 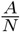 fraction parts are infectious but asymptomatic. 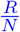 fraction parts are recovered, which are not needed to be isolated due to protective immunity and will remain in the *R* class until the end of the epidemic.

The disease transmission flow chart is depicted in Figure 1 and other parameters are summarized in Table 2. Based on the above assumptions, we formulate the following model to describe the transmission dynamics of COVID-19.

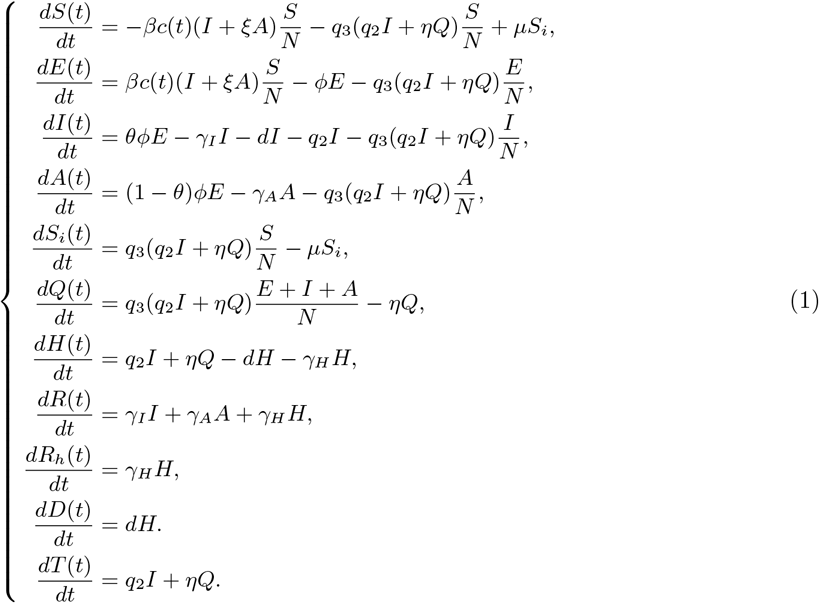

**Table 2.** Definition of the parameters.

| Interpretations | Parameters | Values | Geweke | Source |
| --- | --- | --- | --- | --- |
| Probability of transmission per contact | $\beta$ | 0.054043 | 0.92594 | MCMC |
| Initial contact rate | $c_0$ | 40.319 | 0.99203 | MCMC |
| Transition rate of exposed individuals to the infected class | $\phi$ | 1/5.2 | – | [6] |
| Probability of having symptoms among infected individuals | $\theta$ | 0.6628 | 0.91799 | MCMC |
| Transition rate of quarantined infected individuals to hospital class | $\eta$ | 17.379 | 0.59129 | MCMC |
| Recovery rate of symptomatic infectious individuals | $\gamma_I$ | 0.15796 | 0.7351 | MCMC |
| Recovery rate of asymptomatic infected individuals | $\gamma_A$ | 0.55671 | 0.99168 | MCMC |
| Recovery rate of quarantined infected individuals | $\gamma_H$ | 0.035352 | 0.97782 | MCMC |
| Disease-induced death rate | $d$ | $5.5888 \times 10^{-4}$ | 0.98847 | MCMC |
| Correction factor of transmission probability with asymptomatic infectious individuals | $\xi$ | 0.80987 | 0.74505 | MCMC |
| Rate at which the quarantined uninfected contacts were released | $\mu$ | 1/14 | – | [2] |
| Intervention coefficient with respect to contact | $q_1$ | – | – | Assumption |
| Intervention parameter with respect to patient detection | $q_2$ | 0.47218 | 0.9302 | MCMC |
| Intervention parameter with respect to close contact tracing | $q_3$ | 2.6954 | 0.55476 | MCMC |
| Exponential decreasing rate of contact rate | $\delta$ | $2.8286 \times 10^{-4}$ | 0.99962 | MCMC |

**Figure 1.**
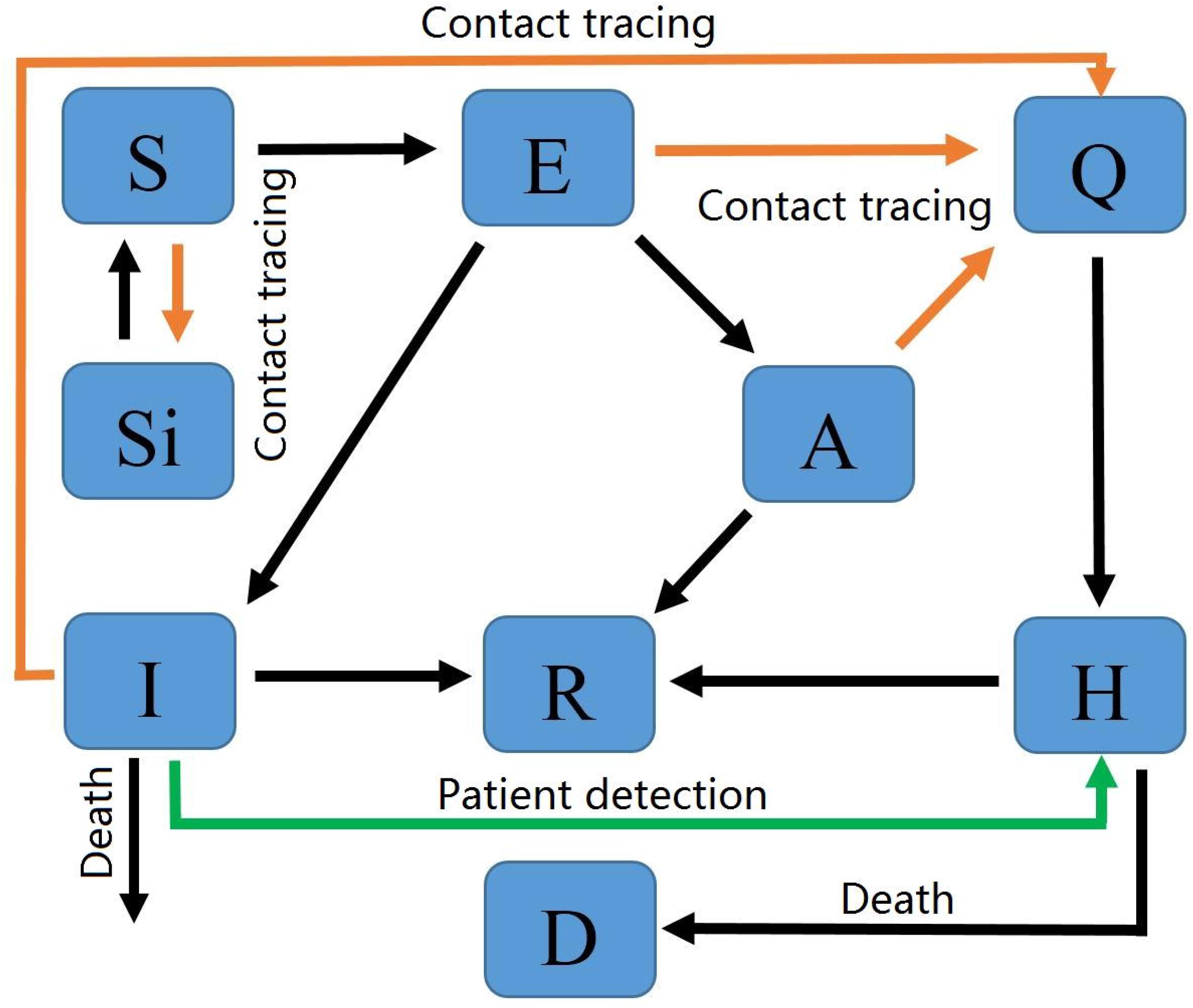
Flow chart of COVID-19 with intervention measures.

According to the concept of next generation matrix in [20] and the basic reproduction number presented in [21], we calculate the basic reproduction number with control measures, i.e. the control reproduction number, *R*_*c*_, of COVID-19, which is given by

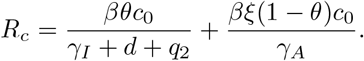

With the spreading of the COVID-19, increasingly intensive intervention measures have been implemented and people gradually enhanced self-protection. In order to quantity the daily reproduction number, inspired by Tang et al. [13], the initial contact rate *c*_0_ in the formula of *R*_*c*_ is replaced by the aforementioned time-dependent contact rate *c*(*t*) to reflect the changes of intervention measures and people’s behaviors. Thereby, we define

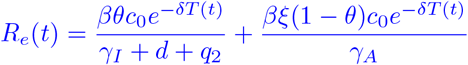

as the effective daily reproduction ratio, the average number of new infections induced by a single infected individual during the infectious period at time *t*. The basic reproduction number or the control reproduction number, which is not time-dependent, can depict the transmission risk in the early phase of disease transmission. While the time-dependent effective daily reproduction ratio can evaluate the transmission risk changing over time.

### Data

COVID-19 daily data excluding Hubei province were archived from NHCC from January 20th (the first day that the number of confirmed cases was reported) to March 3rd, 2020, as shown in Figure 2 ([2]). The data include the cumulative confirmed cases, the cumulative number of deaths, newly confirmed cases and the cumulative number of recovered cases, by reporting date. The data from January 20th to February 24th were fitted to parameterize the model and the data from February 25th to March 3rd were used for comparison of the predicted curves and real data.

**Figure 2.**
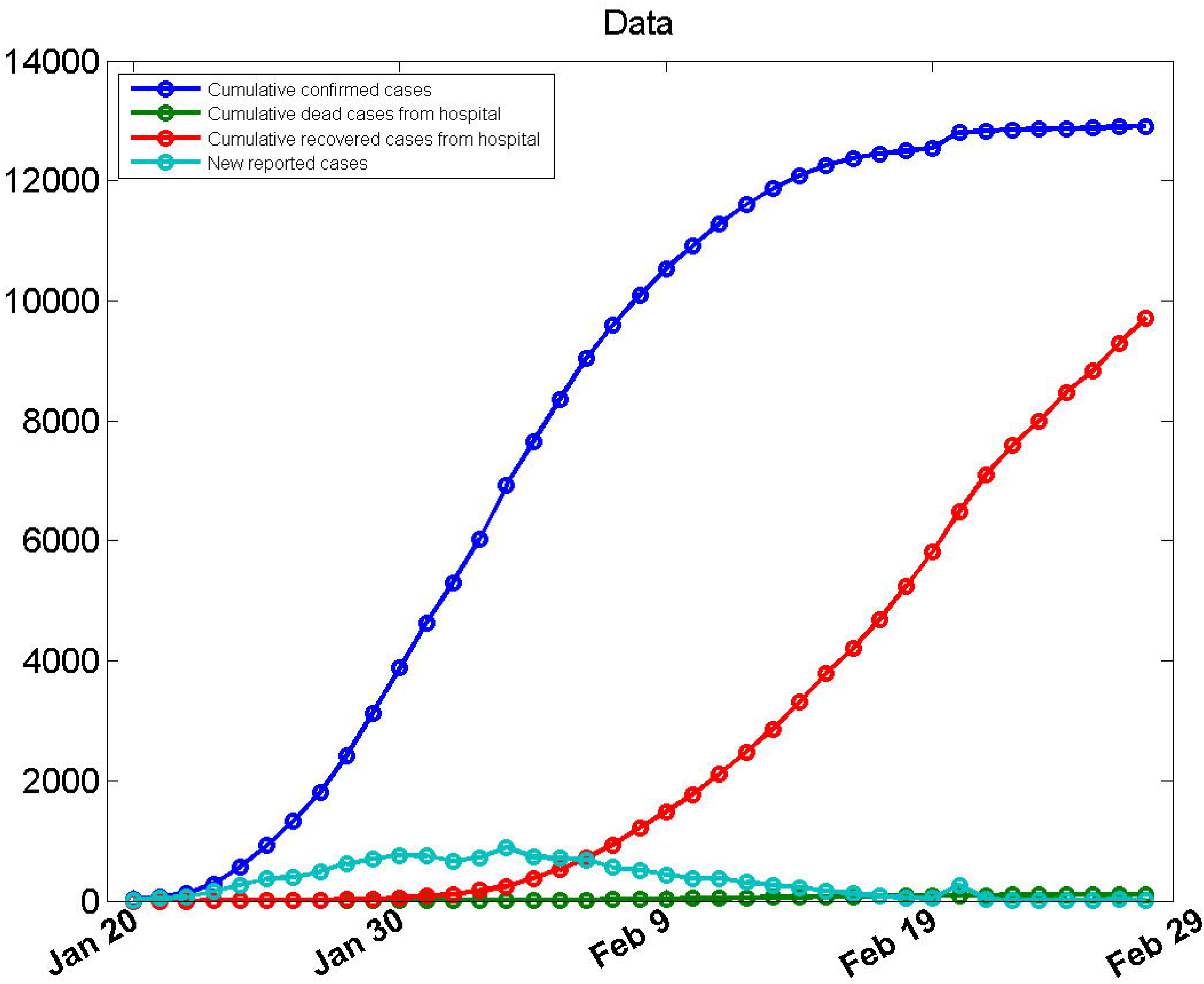
Reported data of mainland China excluding Hubei.

### Parameter estimation

We used the Markov Chain Monte Carlo (MCMC) method with an adaptive Metropolis-Hastings (M-H) algorithm to fit the model. The M-H algorithm, a powerful Markov chain method to simulate multivariate distributions, was executed by using the MCMC toolbox introduced in [23]. This toolbox provides tools to generate and analyse Metropolis-Hastings MCMC chains using multivariate Gaussian proposal distribution. The covariance matrix of the proposal distribution can be adapted during the simulation according to adaptive schemes described in [24, 25, 26].

The algorithm is run for 110,000 iterations with a burn-in of the first 80,000 iterations, and the Geweke convergence diagnostic method is employed to assess the convergence of chains. At the significance level of 5%(the critical value of *z* is 1.96), all parameters and initial values estimated do not reject the original hypothesis of convergence to a posterior distribution (Figure 3).

**Figure 3.**
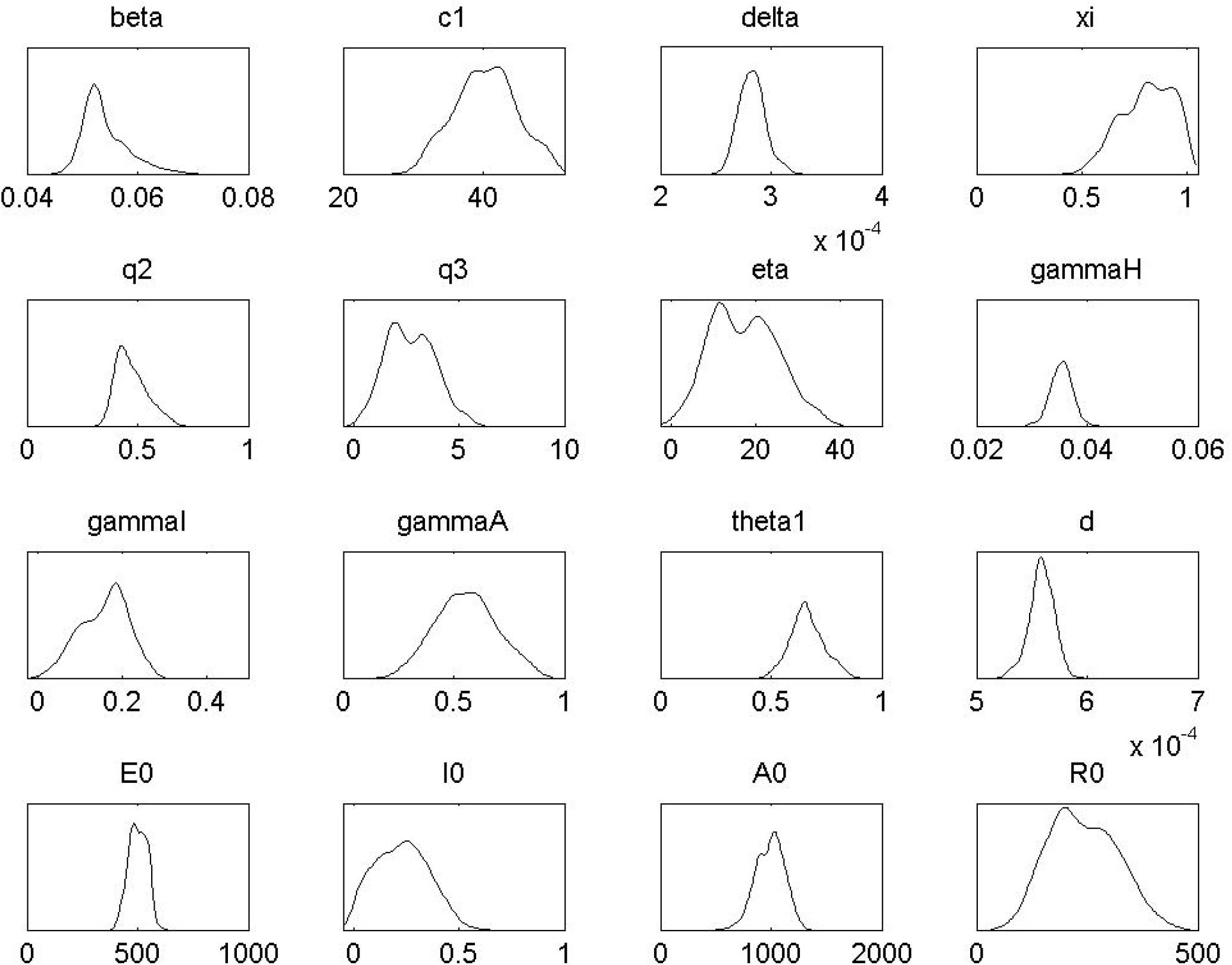
Posterior distributions of each parameter. x-axis is the parameter value, y-axis is the density.

### Simulations

The population of mainland China excluding Hubei province is around 1,336,210,000 [27], which can be set to be the value of *S*(0). 21 confirmed cases were reported on January 20th, 2020 for the first time and the numbers of recovered and dead individuals were both 0. It can be assumed that nobody has been traced by contact tracing at the beginning. Therefore, we set *S*_*i*_(0) = *Q*(0) = *R*_*h*_(0) = *D*(0) = 0, and *H*(0) = *T* (0) = 21. The quarantined susceptible individuals were isolated for 14 days, thus *µ* = 1*/*14 [2]. According to [6], the incubation period of COVID-19 is about 5.2 days, thus *ϕ* can be set to be 1/5.2.

## Results

Figure 4 show that our model yields a relatively good visual fit to the epidemic curves. The MCMC estimation results of each parameters and initial values of some state variables are given in Table 1 and Table 2. By fitting the data, we estimated the control reproduction number*R*_*c*_ to be 3.36 (95% CI 3.20-3.64).

**Figure 4.**
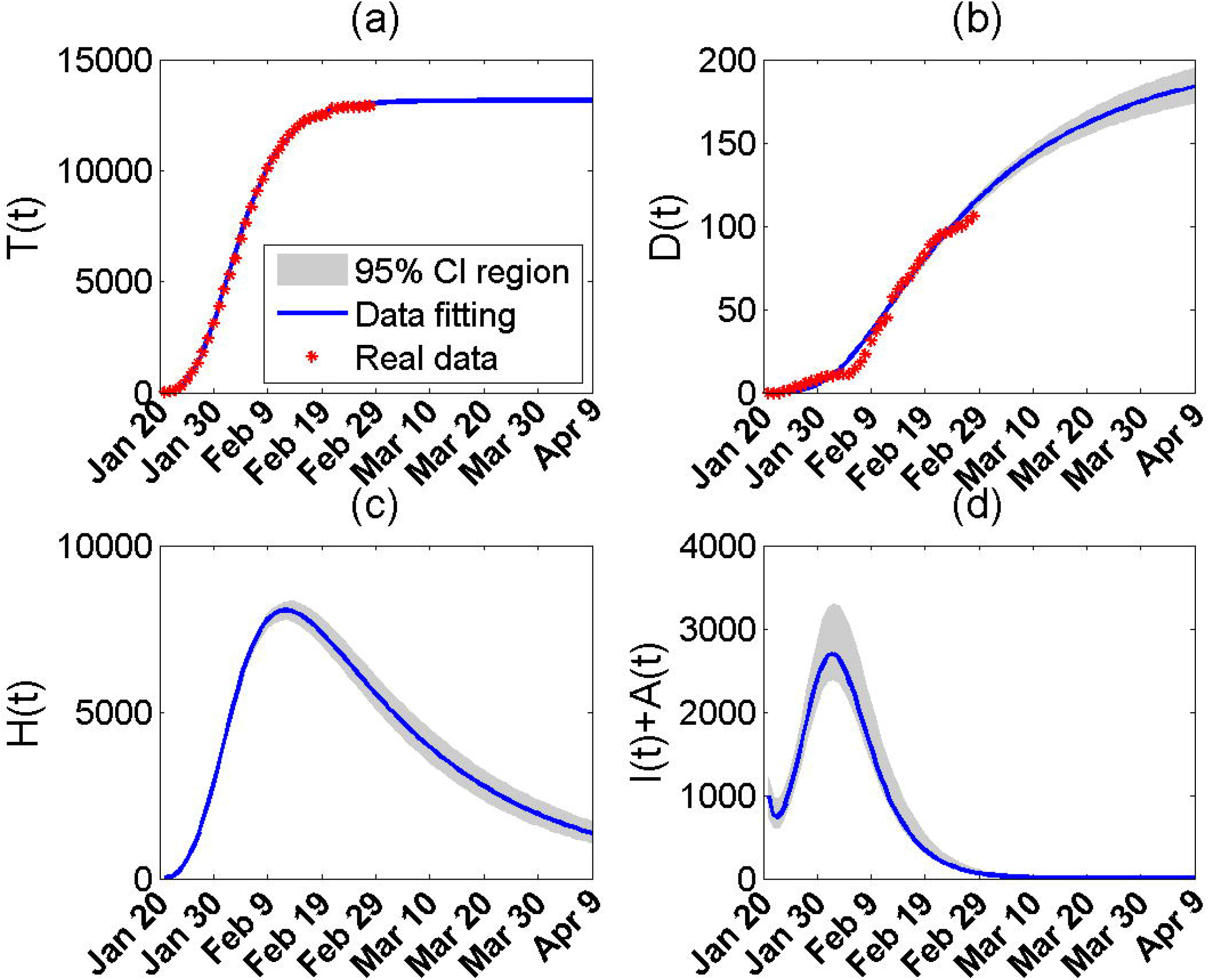
Data fitting and prediction. The abscissa axis is the time. The shaded area is the 95% confidence interval region; The blue solid line is the result of the date fitting; The red stars are the real data. (a): Cumulative confirmed cases; (b): Cumulative dead cases from hospitals; (c): Cumulative recovered cases; (d): The number of New cases; (e): The number of hospitalized individuals; (f): The number of infectious individuals.

Using the estimated parameter values and the number of cumulative cases *T* (*t*), the effective daily reproduction ratio *R*_*e*_(*t*) can be calculated (Figure 5). The result demonstrates that *R*_*e*_(*t*) has dropped sharply from 3.34 (*R*_*e*_(1) = 3.34) on January 20th to 0.89 (*R*_*e*_(12) = 0.89, less than 1) on January 31st, 2020, which implies that the integrated control strategies implemented in mainland China excluding Hubei has successfully reduced transmission intensity and prevented the epidemic growth in a short time frame.

**Figure 5.**
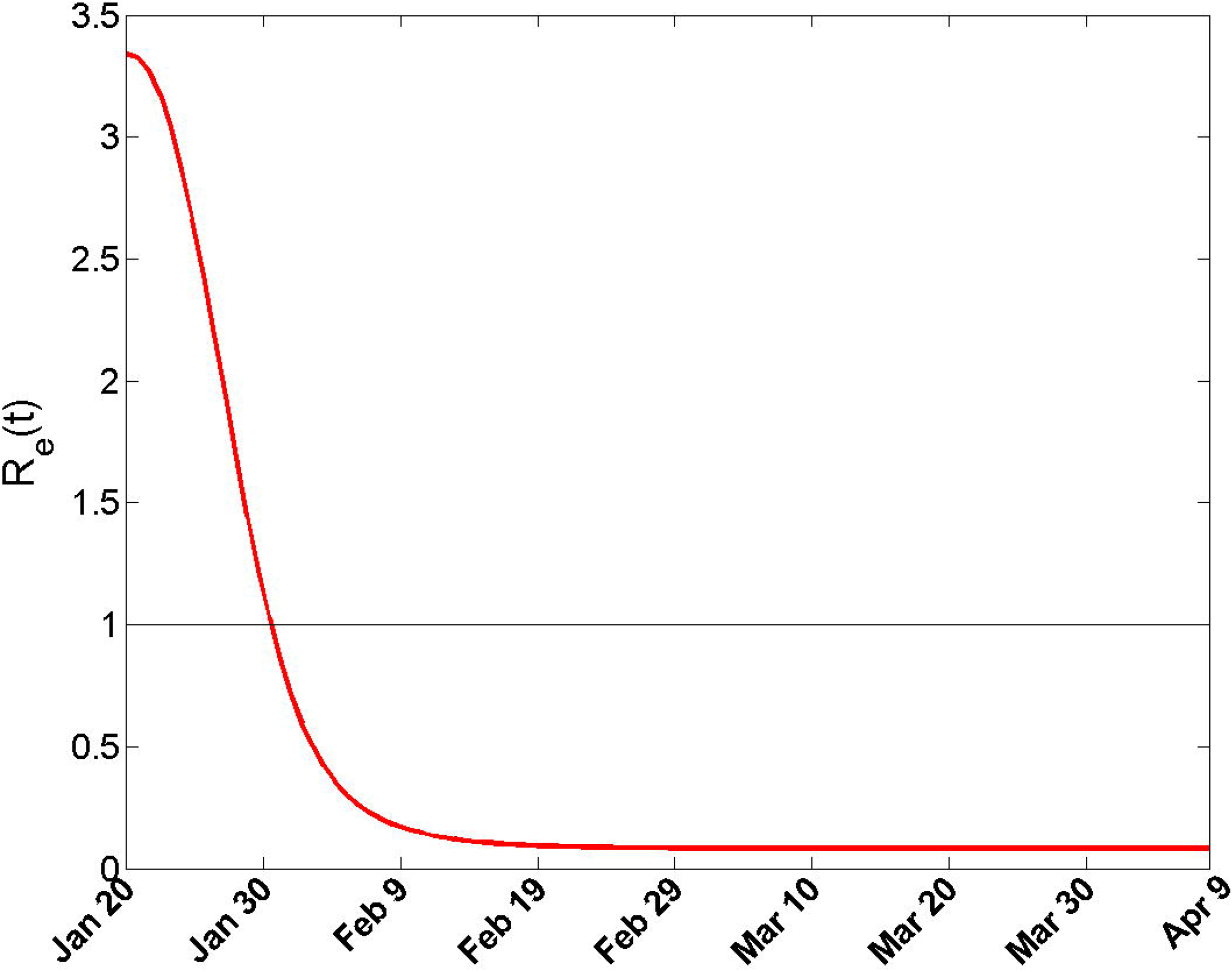
Effective reproduction number.

Under the current rigorous integrated control and self-protection measures, the time series of *T* (*t*) depicted in Figure 4 (a) shows that the cumulative number of confirmed cases will continue growing slowly for some duration and tend to its predicted maximum, which is 13,155. Besides, although the number of hospitalized individuals has peaked on around February 12th, 2020, but it will not shrink to zero in the near future (Figure 4 (e)). Obviously, new infections would occur as long as the infectious individuals who have not been detected exist once people start relieve self-isolation and protection. It is the number of undetected infectious individuals that determines when people’s lives are able to return back to normal. Hence we should closely follow the total number of *I*(*t*) and *R*(*t*). Figure 4 (f) displays that the number of infectious individuals has been decreasing gradually since the end of January. However, it will not descent down to 1 until late March, which infers that people should be fully aware of the real-time epidemic situation and keep personal protect before April.

Using the estimated parameter values and the expression of the effective reproduction ratio *R*_*e*_(*t*), the threshold value of the intervention coefficient with respect to contact, *q*_1_, can be calculated, which is 0.3. This implies that in order to block the continuous spread of the virus, the value of *q*_1_ must be less than 0.3 to guarantee *R*_*e*_(*t*) is below 1. In other words, the contact rate should be kept below 30% of the normal level.

To examine the impact of partial lifting control measures and personal protection, we plot the predicted time series of the number of cumulative confirmed cases, *T* (*t*), and the number of the infectious individuals, *I*(*t*)+*A*(*t*), with different contact rates (Figure 6). Assuming that the adjusting time is from March 5th, 2020, Figure 6 (a) and (b) illustrate that contact rate with 20% of the initial value *c*_0_ will not cause the disease re-bounce. However, the epidemic period will be extended for about 40 days until early May and the cumulative number of confirmed cases will increase by around 0.5% to 13,227. While if the starting time of the adjusting is postponed to March 20th, the epidemic time of disease will be extended for about one week and the cumulative number of confirmed cases will increase by only around 0.05% to 13,161, compared with the scenario of no changes. Nevertheless, if the contact rate is half of the initial value, i.e. *q*_1_ = 0.5 and *R*_*e*_(*t*) = 1.68, COVID-19 will re-bounce on a large scale in a short time frame, even if the starting time is postponed to March 20th. The corresponding analysis results are also listed in Table 3 for comparison.

**Table 3.** The impact of partial lifting control measures and personal protection.

| Starting time of adjustment | Maximum of cumulative confirmed cases | Epidemic period |
| --- | --- | --- |
| No adjustment ($c(t) = c_0 e^{-\delta T(t)}$) | 13,155 | 70 days |
| March 5th ($c(t) = 0.2c_0$) | 13,227 | 110 days |
| March 5th ($c(t) = 0.5c_0$) | More than 447 million | More than 365 days |
| March 20th ($c(t) = 0.2c_0$) | 13,161 | 77 days |
| March 20th ($c(t) = 0.5c_0$) | More than 445 million | More than 365 days |

**Figure 6.**
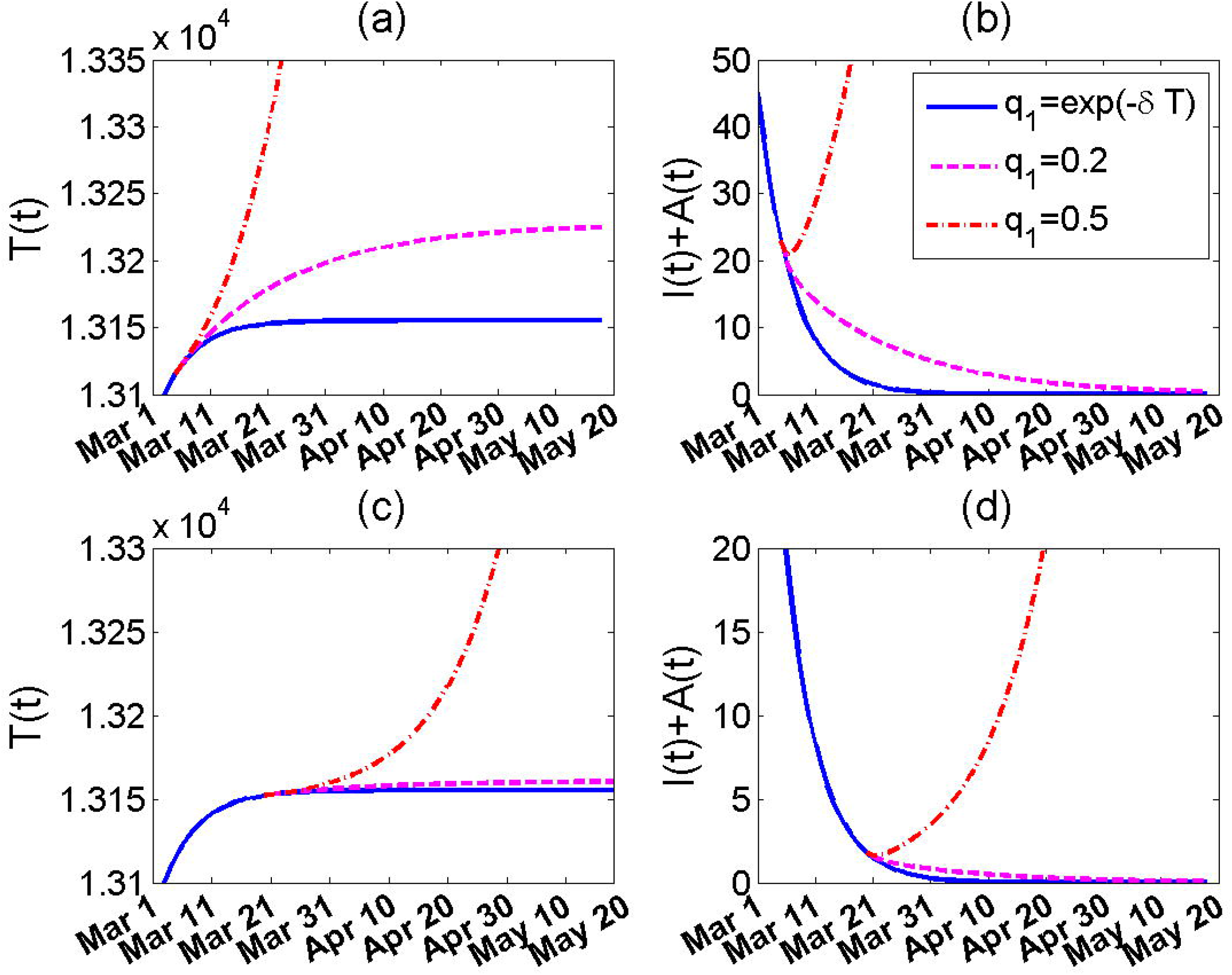
Comparison of scenarios with different contact rates. The abscissa axis is the time. The blue solid line is the case with no changes (*q*_1_ = *exp*(*−δT* (*t*))); The purple dash line is the case with *q*_1_ = 0.2; The red dash-dot line is the case with *q*_1_ = 0.5. (a-b): Adjust the contact rate from March 5th, 2020; (c-d): Adjust the contact rate from March 20th, 2020.

## Discussion

In this paper, we proposed a novel COVID-19 transmission model incorporating the intervention measures implemented in China. Particularly, we adopted two novel function forms which dynamically captured the real-time endemic situation. The first one is the contact rate, *c*(*t*), which was assumed to be dependent on the cumulative number of confirmed cases to better quantify the varied interventions and self-protection measures. The other one is the contact tracing function, which was dependent on the number of new cases.

We evaluated the impact of partial lifting control measures on COVID-19 transmission. Our results show that relieve self-protection too early may lead to the spread of the virus for a longer time and more people would be infected, and may even lead to the mass transmission of the virus again. The reduction of the emergency response level does not mean that people can be off guard. At least until the end of March, life will not be able to return to normal.

In the process of recovery of production and life, we should pay attention to take protective measures to minimize the contact between people, such as wearing masks and trying to avoid crowded places to cut the risk of catching coronavirus. By calculating the effective reproduction ratio, we assert that people’s contact rate should be kept below 30% of the normal level and the lower the better.

The forecasts presented are based on the assumption that there are no imported cases from Hubei and other infected regions. The WHO recently upgraded the global risk of the epidemic to ‘very high’. New endemic foci outside China are formed, such as South Korea, Italy, Iran, Japan, etc. Although the current endemic situation in China is under control, population migration cross country border should be taken into consideration when we modify the updated controlling policy of COVID-19. With the development of the epidemic situation in other countries in the world, it is very important to maintain and strengthen the quarantine of entry personnel. The impact of international mobility on the transmission of COVID-19 will be studied in our future works.

We concentrated on the epidemic situation in mainland China excluding Hubei province is due to the significant differences between Wuhan, Hubei, and the rest of the country. In Wuhan, because of the sudden appearance of the disease, it took much longer time to recognize and understand the disease transmission than other regions. It requires certain time from unknown to known. Therefore, the data accuracy in Wuhan, Hubei is a major issue for parameters’ calibration. In addition, the sudden large outbreak in Wuhan, Hubei exhausted all medical resources in a short time. A more targeted model considering medical resource capacity will be anticipated in the future.

Our results manifest that the model yields a good visual fit to the epidemic curve except for the cumulative recovered cases. The number of predicted recovered cases is less than the number of actual recovered individuals in the later period of data fitting. The possible reason is that the recovery rate of hospitalized individuals was increasing with the improvement of treatment level and the increase of medical resources. In addition, due to increased contact tracing efficiency, large quantity of mild symptomatic cases were detected and hospitalised which has higher recover rate than other classified cases. A time-dependent recovery rate may be more suitable to fit the real data. Furthermore, age is an key parameter determining the mortality and recover rates. It is ideal to set up age-structured agent-based or meta population-based deterministic models which can capture this important risk factor. In this work, the initial value of the susceptible class *S*(0) was set to be the population of mainland China excluding Hubei province. Actually, the number of susceptible individuals is not as many as this number considering the very strict control measures implemented and the difference of behaviors between different age groups. A proper method to estimate the accurate number of susceptible individuals should be studied in the future. Besides, the impact of limited medical resources is another important issue that is not incorporated in our model. The transmission dynamics of COVID-19 accounting for the potential negative effects of health systems being overwhelmed on mortality will be studied in our future work.

## Conclusions

In conclusion, to ensure the COVID-19 epidemic ending rapidly, it is necessary to maintain the current integrated control intervention and self-protection measures, including travel restriction, quarantine of entry, contact tracing followed by quarantine and isolation and reduction of contact, like wearing masks, etc. People should be fully aware of the real-time epidemic situation and keep sufficient personal protection until April. If all the above conditions are met, the outbreak is expected to be ended by April in mainland China apart from Hubei province.

## Data Availability

All data were obtained from the National Health Commission of China.

http://en.nhc.gov.cn/

## Abbreviations

COVID-19: Corona Virus Disease 2019;
NHCC: National Health Commission of the People’s Republic of China;
MCMC: Markov Chain Monte Carlo method;
*R*_0_: Basic reproduction number;
*R*_*c*_: Control reproduction number;
*R*_*e*_(*t*): Effective daily reproduction ratio.

## Author’s contributions

HW designed research; HW conceived the experiments, HW, JC and GY conducted the experiments and analyzed the results; HW and GJ wrote the manuscript. All authors read and approved the final manuscript.

## Acknowledgements

The authors thank graduate students Yuanyuan Yu and Jingyuan Li for their efforts in collecting data during the study.

## Availability of data and materials

Not applicable.

## Ethics approval and consent to participate

Not applicable.

## Consent for publication

Not applicable.

## Tables

section*Tables

section*Tables

## Additional Files

Additional file 1 — Sample additional file title

Additional file descriptions text (including details of how to view the file, if it is in a non-standard format or the file extension). This might refer to a multi-page table or a figure.

Additional file 2 — Sample additional file title

Additional file descriptions text.

